# Visceral Adiposity and Subclinical Left Ventricular Remodeling

**DOI:** 10.1101/2023.11.21.23298826

**Authors:** Judy Luu, Catherine Gebhard, Matthias G. Friedrich, Dipika Desai, Karleen M Schulze, Russell de Souza, Baraa K. Al-Khazraji, Philipp Awadalla, Guillaume Lettre, Vikki Ho, Trevor Dummer, Jason Hicks, Marie-Eve Piche, Paul Poirier, Koon K. Teo, Salim Yusuf, Jean-Claude Tardif, Jennifer Vena, Douglas S. Lee, Francois Marcotte, Eric Larose, Eric E. Smith, Sonia S. Anand, CAHHM Study Investigators

**Author notes:** These authors contributed equally to this work (Judy Luu, Catherine Gebhard). **Corresponding author:** Sonia S Anand Population Health Research Institute Hamilton Health Sciences, McMaster University 237 Barton St East, Hamilton, Ontario L8L 2X2, Canada (Ext. 21523). **Competing interests** The University Hospital Zurich (CG) holds a research contract with GE Healthcare. Matthias G. Friedrich is board member, shareholder, and consultant of Circle Cardiovascular Imaging Inc. **Role of the Funder/Sponsor:** The funding sources had no role in the design and conduct of the study; collection, management, analysis, and interpretation of the data; preparation, review, or approval of the manuscript; and decision to submit the manuscript for publication.

## Abstract

**Introduction:** Visceral adiposity is emerging as a key driver of cardio-metabolic risk factors and cardiovascular disease (CVD), but its relationship with cardiac structure and function is not well characterized across sexes. Using the Canadian Alliance for Healthy Heart and Minds (CAHHM), a large population-based cohort study, we sought to determine the association of visceral adipose tissue (VAT) on subclinical left ventricular (LV) remodeling in males and females.

**Methods:** As part of the CAHHM study, 6522 participants free of clinical CVD (mean age: 57.4 [8.8 SD] years; 3,671 females, 56%) underwent magnetic resonance imaging (MRI) in which LV parameters and VAT volume were measured. Information about demographic factors, CV risk factors, and anthropometric measurements were obtained. Subclinical cardiac remodelling was defined as altered LV concentricity, represented by increased LV mass-to-volume ratio (LVMV).

**Results:** Males had a higher VAT volume (80.8 mL; 95% CI: 74.6 t 86.9) compared to females (64.7 mL; 95% CI: 58.5 to 70.8), adjusted for age and height. Among both males and females, VAT was significantly associated with subclinical cardiac remodeling (increased LVMV), independent of other CV risk factors. In multiple regression models adjusted for cardiovascular risk factors, age, and height, every 1 sex-specific standard deviation increase in VAT corresponded to an increase of 0.037 g/mL in LVMV (95% CI: 0.032 to 0.041; p<0.001), which was consistent across both sexes. Notably, a 1 standard deviation increase in VAT is associated with a LVMV that is 20 times higher than what is observed with natural aging alone (0.0020 g/mL rise in LVMV (95% CI 0.0016 to 0.0025), and 1.5 times higher than the impact of an integrated measure of CV risk factors (0.024 g/mL; 95% CI: 0.020 to 0.028).

**Conclusion:** VAT significantly influences subclinical cardiac remodeling in both males and females, independent of other cardiovascular risk factors and age. Further research to understand the pathways by which VAT contributes to accelerated cardiac aging is needed.

## Introduction

Over the last several decades, population-based cohort studies have shown that subclinical cardiovascular disease (CVD) represent asymptomatic stages that can progress to symptomatic CVD,^1,2,3^ These studies, including the Multi-Ethnic Study of Atherosclerosis (MESA) and the Framingham Study, have previously reported that geometric changes of the left ventricle (LV) (i.e., remodeling) precede adverse cardiovascular (CV) outcomes.^1,2^ Specifically, increased concentricity (relationship between wall thickness and cavity size), defined by the LV mass to volume ratio (LVMV), is associated with 2-fold increased risk for coronary heart disease and 4-fold risk for stroke.^1,2^ Moreover, biological aging is identified as a risk factor for LV remodeling. Previous studies have demonstrated that LVMV increases by 0.005g/mL with each year of age. This change in LVMV, indicative of a hypertrophic process in the setting of progressively reduced LV volume, conferred a greater risk for total cardiovascular events, especially when present earlier in life.^3^

Recently, visceral adipose tissue (VAT) was reported to be associated with increased LVMV in the MESA cohort, independent of age, sex, race, or obesity status.^5^ This study, however, used simplified cardiovascular magnetic resonance imaging (CMR) contouring methodology to derive LV mass.^5^ As a component of total body fat, VAT releases different bioactive molecules and hormones, and is directly linked to impaired glucose and lipid metabolism.^7,8^ VAT accumulation further leads to increased susceptibility for arterial hypertension, ischemic heart disease, and cognitive dysfunction.^7–9^ Studies have demonstrated that VAT drives the aging process in the heart by producing inflammatory molecules^10^, profibrotic factors, such as osteopontin, and transforming growth factor β (TGF-β), which promotes interstitial fibrosis and may lead to the development of heart failure.^11,12^

We recently published sex and age-specific normal reference values for CMR parameters derived from the largest, multi-ethnic population free of CVD to date.^6^ We reported accurate normal reference values for LVMV for both males and females, across different age groups, using anatomically-precise contouring methodology that determines LV mass in end-systole and incorporates papillary muscles and trabeculations as part of LV mass. Building on the strength of this large population-based cohort,^13^ we investigated the relationship of VAT on left ventricular remodeling using sex-specific normal reference values for LVMV as an index for LV concentricity.

## Methods

### Study Population

The rationale, study design, site-specific patient characteristics, and follow-up durations of the Canadian Alliance for Healthy Hearts and Minds (CAHHM) cohort have previously been described.^13^ Briefly, the CAHHM study is a large prospective, multi-center cohort study that aims to investigate the contextual, health behaviours, and socio-cultural factors with subclinical CV health as detected by magnetic resonance imaging (MRI). Detailed phenotyping including multi-organ MRI in an ethnically and geographically diverse population was utilized. CAHHM enrolled a total of 8,580 individuals (54% females) between the ages of 30 and 78 years who were recruited from 8 Canadian cohorts **(Supplementary Figure 1).** The study complied with the Declaration of Helsinki, and each study site received institutional research ethics board approval for all procedures. All study participants provided written informed consent.

### Demographic and Lifestyle Data

Standardized questionnaires were used to gather information on demographic variables, including CV risk factors (hypertension, diabetes, smoking, dyslipidemia, family history of coronary artery disease) from 2014to 2018. Hypertension was defined as systolic blood pressure (SBP) ≥140 mmHg or diastolic blood pressure (DBP) ≥90 mmHg, or self-reported use of anti-hypertensive medication. Diabetes mellitus was defined as self-reported history of diabetes with use of diabetes medications. Dyslipidemia was defined when individuals were prescribed use of a lipid-lowering treatment. History of tobacco use was captured as current, former smoker (quit more than 12 months ago), as well as never smoked. All participants underwent physical assessment using a standardized protocol by trained study personnel, including body weight, height, blood pressure, heart rate, waist and hip circumference, and percent body fat measurements (using bioelectrical impedance analysis). Body surface area (BSA) and body mass index (BMI) were calculated using height and weight measurements. Resting heart rate was obtained after resting seated for 10 minutes.

Cardiovascular risk factor burden were summarized by the INTERHEART^14^ and the modified Framingham Risk Scores (FRS).^15,16^ Similar to the FRS, the non-laboratory-based INTERHEART Risk Score (IHRS) is a validated score that reflects the quantified CV risk factor burden, and is predictive of subclinical and clinical CVD.^14,17^ The IHRS includes age, sex, smoking status, diabetes, high blood pressure, family history of MI, waist-to-hip ratio, home or work social stress, depression, dietary habits, and physical activity. The IHRS scores range from 0 to 48; where low risk is defined as a score of 0 to 9, moderate risk as 10 to 16, and high risk as 17 or higher.

Ethnicity or race was based on self-report using the following categories: Indigenous, Arab, Black, East Asian, Filipino, South Asian, Jewish, Latin American/Hispanic, Southeast Asian, West Asian, White European descent, and other ethnic groups not listed (free text answer). In Table 1, the ethnicity categories were confined to the four largest groups (White European, South Asian, East Asian, Southeast Asian) and reported accordingly.

**Table 1:** Baseline demographics, medical history, and anthropometric measurements. Presented data are n (%) unless otherwise specified. ^a^Includes self-identified Black individuals, Indigenous people, Mixed and unknown ethnicity. ^b^Framingham Risk score modified to use Apolipoprotein B and A-1 as approximation for LDL and HDL, respectively.

|  | **N** | **Overall** | **Females** | **Males** |
| --- | --- | --- | --- | --- |
| Number of participants | 6522 | 6522 | 3671 | 2851 |
| Age, mean (SD), y | 6522 | 57.4 (8.8) | 56.9 (8.6) | 58.0 (9.0) |
| **Self-reported ethnicity** |  |  |  |  |
| East & South East Asian | 6522 | 898 (13.8) | 512 (13.9) | 386 (13.5) |
| South Asian | 6522 | 230 (3.5) | 100 (2.7) | 130 (4.6) |
| White European | 6522 | 5262 (80.7) | 2976 (81.1) | 2286 (80.2) |
| Other<sup>a</sup> | 6522 | 132 (2.0) | 83 (2.3) | 49 (1.7) |
| **Smoke status** |  |  |  |  |
| Current | 6522 | 337 (5.2) | 183 (5.0) | 154 (5.4) |
| Former | 6522 | 2194 (33.6) | 1208 (32.9) | 986 (34.6) |
| Never | 6522 | 3991 (61.2) | 2280 (62.1) | 1711 (60.0) |
| **Medical History** |  |  |  |  |
| Diabetes on treatment | 6519 | 283 (4.3) | 116 (3.2) | 167 (5.9) |
| Elevated cholesterol &/or regular use of statins | 6522 | 2349 (36.0) | 1064 (29.0) | 1285 (45.1) |
| Hypertension | 6521 | 2411 (37.0) | 1063 (29.0) | 1348 (47.3) |
| **INTERHEART risk score** | 6522 | 9.9 (5.7) | 8.7 (5.3) | 11.4 (5.7) |
| Low | 6522 | 3381 (51.8) | 2231 (60.8) | 1150 (40.3) |
| Moderate | 6522 | 2074 (31.8) | 1029 (28.0) | 1045 (36.7) |
| High | 6522 | 1067 (16.4) | 411 (11.2) | 656 (23.0) |
| **Framingham risk score<sup>b</sup>** | 3339 | 11.6 (4.1) | 10.9 (4.0) | 12.4 (3.9) |
| Low | 3339 | 1682 (50.4) | 1174 (66.6) | 508 (32.2) |
| Moderate | 3339 | 1086 (32.5) | 477 (27.1) | 609 (38.6) |
| High | 3339 | 571 (17.1) | 112 (6.4) | 459 (29.1) |
| **Physical Measures** |  |  |  |  |
| Weight, kg, mean (SD) | 6522 | 75.8 (16.2) | 69.7 (15.0) | 83.7 (14.1) |
| Height, cm, mean (SD) | 6522 | 168.3 (9.3) | 162.7 (6.6) | 175.6 (7.0) |
| Body Mass Index, mean (SD), kg/m<sup>2</sup> | 6522 | 26.7 (4.8) | 26.3 (5.3) | 27.1 (4.1) |
| BMI, kg/m<sup>2</sup>, mean (SD) |  |  |  |  |
| <25 (Normal) | 6522 | 2655 (40.7) | 1737 (47.3) | 918 (32.2) |
| 25-29 (Overweight) | 6522 | 2505 (38.4) | 1152 (31.4) | 1353 (47.5) |
| 30-34 (Obese) | 6522 | 953 (14.6) | 500 (13.6) | 453 (15.9) |
| 35+ (Severely Obese) | 6522 | 409 (6.3) | 282 (7.7) | 127 (4.5) |
| Percent Body Fat, %, mean (SD) | 6502 | 30.5 (9.1) | 35.1 (8.1) | 24.6 (6.8) |
| Waist, cm, mean (SD) | 6522 | 88.2 (13.6) | 83.8 (13.4) | 93.8 (11.7) |
| Hip, cm, mean (SD) | 6522 | 101.0 (10.6) | 101.0 (12.1) | 101.1 (8.3) |
| Waist to hip ratio, mean (SD) | 6522 | 0.87 (0.08) | 0.83 (0.07) | 0.93 (0.07) |
| Visceral Adipose Tissue, mL,<br>mean (SD) | 6522 | 71.0 (36.5) | 61.3 (30.3) | 83.6 (39.9) |

### Magnetic Resonance Imaging Protocol and Image Acquisition

All CAHHM study participants underwent a multi-level MRI for the heart, neck and head, performed at a field strength of 1.5T or 3.0T between 2013 and 2018. Images were obtained according to a standardized protocol.^13^ Briefly, cine CMR images were acquired using a single-breath hold, retrospective electrocardiogram (ECG)-gated steady state free precession (SSFP) sequence in both the long axis (2 slices) and short axis (SAX) views (12-14 slices). Field of view was 360mm, voxel size ranged from 0.9 to 1.2mm, while slice thickness was 8 mm, with a 2mm gap.

### Visceral Adipose Tissue

Abdominal VAT was determined by T1-weighted MRI sequences, providing a bright signal for adipose tissue, and specifically via T1-weighted turbo spin echo axial sequence through the lumbar, L4-L5, levels as previously described.^9^ VAT volumes were quantified in mL and analyzed/reported by the core laboratory, with sex-stratified quartiles derived for analysis.

### Cardiac MRI Analysis

Images were stored by using the infrastructure of the Canadian Atherosclerosis Imaging Network (CAIN). Image analysis was performed by core lab experts at four different institutions (Montreal Heart Institute, Montreal; Sunnybrook Health Sciences Centre, Toronto; Institut Universitaire de Pneumologie et Cardiologie, Québec City; University of Calgary, Calgary). Analysis of CMR images was performed at the Montreal Heart Institute, Montreal using certified software (cvi42, Circle CV Imaging Inc., Calgary, Canada). LV mass, volumes, and functional parameters were determined from contiguous short axis cine images covering the heart from base to apex throughout the cardiac cycle, with a temporal resolution of less than or equal to 50ms, as previously described.^13^ LV end-diastolic volume (LVEDV) and end-systolic volume (LVESV) were determined by summing myocardial areas on each separate slice multiplied by the sum of the slice thickness and image gap (slice summation method) with interpolated gap data. LV mass was calculated by summation of the myocardial area multiplied by slice thickness plus image gap in the end-systolic phase multiplied by the myocardial specific gravity (1.05 g/mL).^18^ Anatomical accuracy was verified by carefully tracing contours for LV mass at end-systole, including papillary muscles and trabecular tissue into LV mass.^6^ LV mass was determined from the end-systolic phase due to several reasons: the endocardial border line is shorter, minimizing error risk; systole has fewer slices than diastole, leading to fewer contours and potential mistakes; and closed inter-trabecular recesses in systole reduce the likelihood of partial volume errors.^6^ LV stroke volume (LVSV) was derived by subtracting LVESV from LVEDV. LV ejection fraction (EF) was calculated by dividing LVSV with LVEDV and multiplying by 100. Cardiac output was derived as LVSV×heart rate. LV mass-to-volume ratio (LVMV) was calculated by dividing LV systolic mass by LVEDV. Volumetric parameters were indexed to BSA.

### Statistical Analysis

In this cross-sectional analysis, demographic and lifestyle descriptors are presented overall and by sex (Table 1). Generalized linear mixed models (GLMM) with centre modelled with random intercepts and an unstructured covariance matrix were used to describe LV measures and VAT overall and testing differences between males and females, adjusting for age and ethnicity. Similar GLMM were used to describe CV risk factors and CMR variables over sex-specific VAT quartiles, stratified by sex. Variables were tested for trend over the VAT quartiles using linear contrasts. To account for sex-specific variations in VAT, a standardized VAT value was introduced, determined as [(VAT-mean)/sex-specific standard deviation]. A final multivariable GLMM was used to estimate the effect of the standardized VAT on LVMV adjusting for age, sex, height, the INTERHEART risk score and ethnicity (not shown). Sex stratified models and testing interactions supported the decision to combine males and females for these analyses. All analyses were conducted in SAS 9.4 (SAS Institute, Cary, NC).

## Results

### Patient Characteristics and CMR Parameters

A total of 8,580 participants consented for the study with MRI data available for 8,258 subjects. Due to history of CVD (873), missing LV parameters/poor image quality (350), or missing VAT or IHRS variables (513), 1736 subjects were excluded **(Suppl Figure 1).** Therefore, a total of 6,522 participants with CMR examinations were included in the analysis (3,671 females, 56%). The mean age was 58±9 years for males and 57±9 years for females.

Details of the baseline demographics for the males and females are shown in **Table 1**. Overall, there was an even distribution of males and females across the different ethnicities. Most subjects were of White European origin; approximately 14% were of East and Southeast Asian ethnicity, and approximately 4% were of South Asian ethnicity. The burden of CV risk factors was greater in males than females, with 61% of males having moderate to high IHRS scores, compared to 40% of females **(Table 1). Table 2** shows the estimated mean LV parameters, including LVMV, in the overall population and in each sex.

**Table 2:**
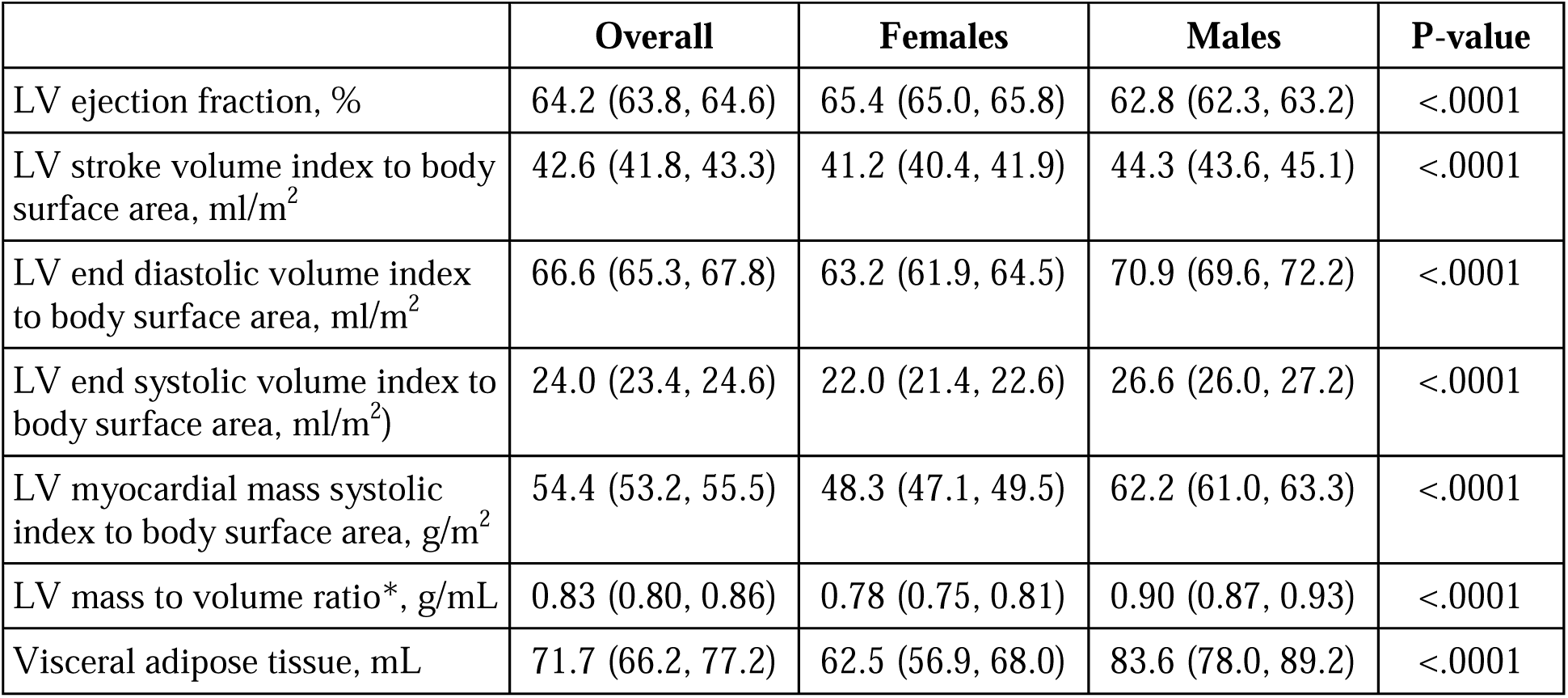
Left ventricular Cardiac magnetic resonance imaging parameters. Presented data are means (95% CI) adjusted for age, (sex) and ethnicity with fixed effects and centre with random intercepts. *Mass to volume ratio derived from LV myocardial mass in systole (g) divided by LV end diastolic volume. Abbreviations: LV, left ventricular.

|  | **Overall** | **Females** | **Males** | **P-value** |
| --- | --- | --- | --- | --- |
| LV ejection fraction, % | 64.2 (63.8, 64.6) | 65.4 (65.0, 65.8) | 62.8 (62.3, 63.2) | <.0001 |
| LV stroke volume index to body surface area, ml/m<sup>2</sup> | 42.6 (41.8, 43.3) | 41.2 (40.4, 41.9) | 44.3 (43.6, 45.1) | <.0001 |
| LV end diastolic volume index to body surface area, ml/m<sup>2</sup> | 66.6 (65.3, 67.8) | 63.2 (61.9, 64.5) | 70.9 (69.6, 72.2) | <.0001 |
| LV end systolic volume index to body surface area, ml/m<sup>2</sup>) | 24.0 (23.4, 24.6) | 22.0 (21.4, 22.6) | 26.6 (26.0, 27.2) | <.0001 |
| LV myocardial mass systolic index to body surface area, g/m<sup>2</sup> | 54.4 (53.2, 55.5) | 48.3 (47.1, 49.5) | 62.2 (61.0, 63.3) | <.0001 |
| LV mass to volume ratio*, g/mL | 0.83 (0.80, 0.86) | 0.78 (0.75, 0.81) | 0.90 (0.87, 0.93) | <.0001 |
| Visceral adipose tissue, mL | 71.7 (66.2, 77.2) | 62.5 (56.9, 68.0) | 83.6 (78.0, 89.2) | <.0001 |

### Sex Differences in the Relationship between VAT with LV Parameters

Adjusted for age, height, and ethnicity, females had a higher percentage body fat (35.6%; 95% CI: 35.0-36.3) compared with males (24.5%; 95% CI: 23.8, 25.2), whereas males had significantly more VAT (80.8 mL; 95% CI: 74.6, 86.9) than females (64.7 mL; 95% CI: 58.5, 70.8).

**Tables 3a and 3b** show the VAT by quartile for females and males, respectively. In general, higher INTERHEART risk scores and the presence of CV risk factors were associated with higher VAT. Similarly, higher height and percent body fat was associated with higher VAT. In each successive VAT quartile, the LVMV was higher, driven predominantly by decreases in LVEDV in females, and both decreases in LV mass and LVEDV in males.

**Table 3a:**
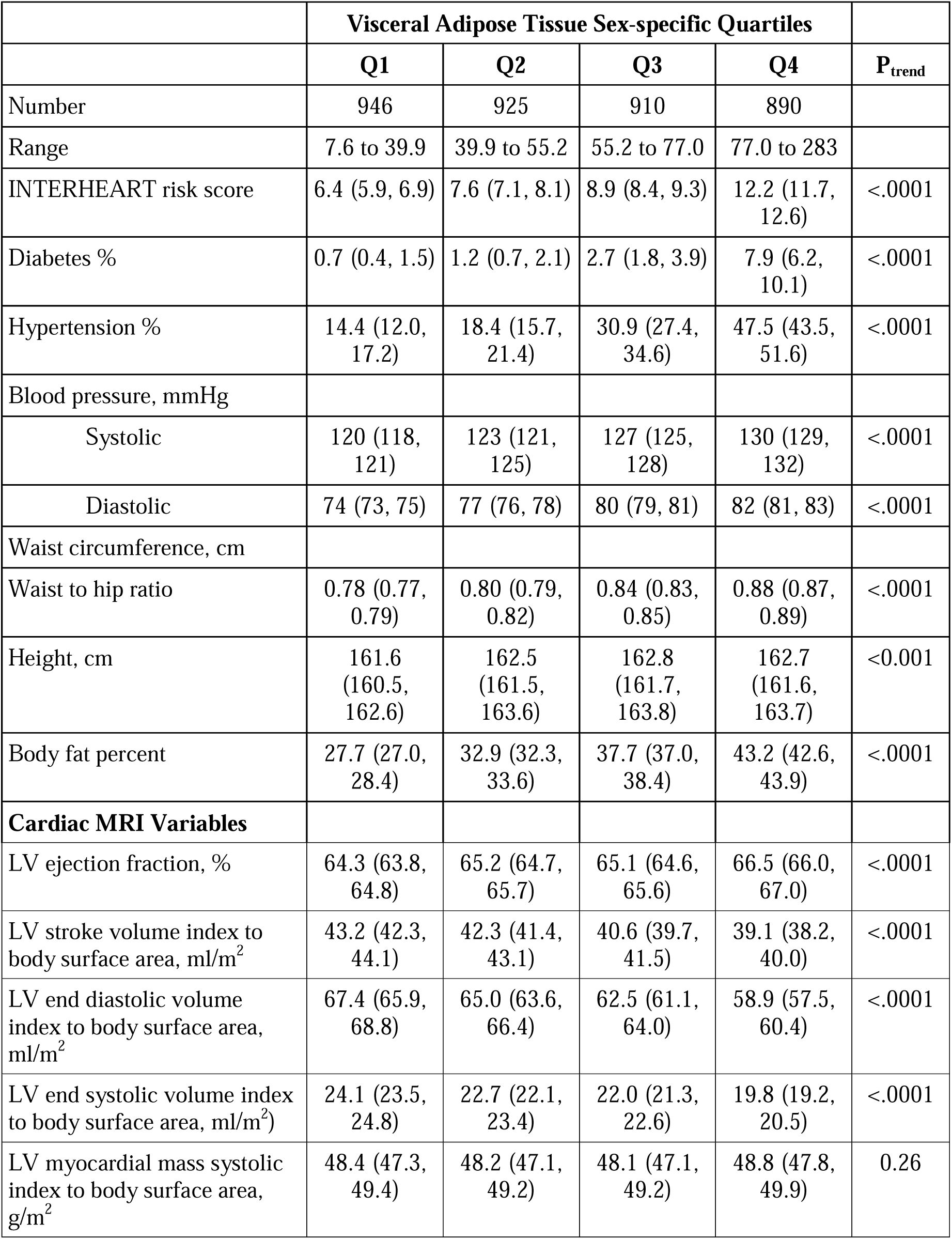
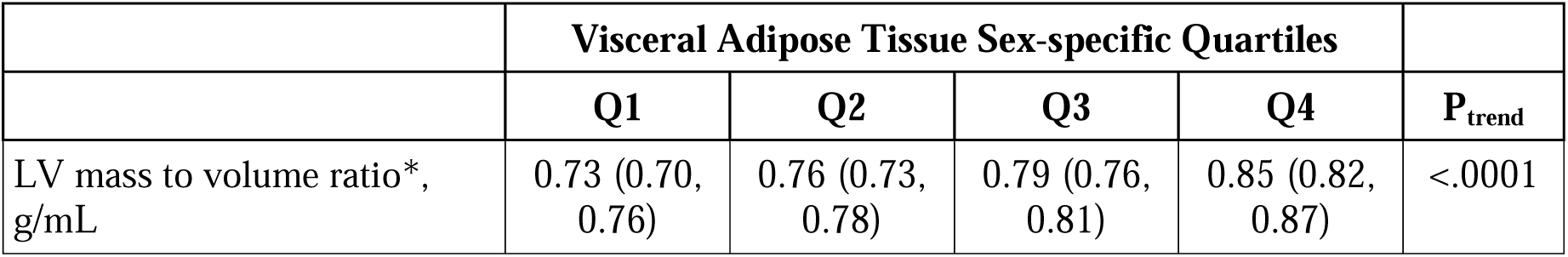
Cardiovascular risk profiles and cardiac magnetic resonance imaging parameters across different visceral adipose tissue quartiles for females. Data presented are adjusted means or proportions (95% CI) from generalized linear mixed models. Fixed effects include age and ethnicity and centre modelled as a random effect. P-value for trend calculated using linear contrasts. Diabetes includes participants with any type (including type1 and type 2 diabetes) taking medication for diabetes; Hypertension includes those on blood pressure reducing medication or with an elevated blood pressure reading as determined by systolic blood pressure >140 mmHg or diastolic blood pressure >90 mmHg.

**Table 3b:** Cardiovascular risk profiles and cardiac magnetic resonance imaging parameters across different visceral adipose tissue quartiles for males. Data presented are adjusted means or proportions (95% CI) from generalized linear mixed models. Fixed effects include age, and ethnicity and centre modelled as a random effect. P-value for trend calculated using linear contrasts. Diabetes includes participants with any type (including type1 and type 2 diabetes) taking medication for diabetes; Hypertension includes those on blood pressure reducing medication or with an elevated blood pressure reading as determined by systolic blood pressure >140 mmHg or diastolic blood pressure >90 mmHg.

|  | Visceral Adipose Tissue Sex-specific Quartiles |  |  |  |  |
| --- | --- | --- | --- | --- | --- |
|  | Q1 | Q2 | Q3 | Q4 | P<sub>trend</sub> |
| Number | 730 | 724 | 698 | 699 |  |
| Range | 11.4 to 54.8 | 54.8 to 77.6 | 77.6 to 106 | 106 to 303 |  |
| INTERHEART risk score | 8.8 (8.2, 9.4) | 10.6 (10.0, 11.1) | 12.6 (12.1, 13.2) | 15.1 (14.5, 15.7) | <.0001 |
| Diabetes % | 2.0 (1.2, 3.3) | 2.9 (1.9, 4.4) | 5.3 (3.9, 7.3) | 11.7 (9.4, 14.5) | <.0001 |
| Hypertension % | 30.6 (26.8, 34.5) | 41.7 (37.7, 45.9) | 51.8 (47.5, 56.0) | 66.4 (62.2, 70.4) | <.0001 |
| Blood pressure, mmHg |  |  |  |  |  |
| Systolic | 130 (129, 132) | 133 (132, 135) | 135 (133, 137) | 138 (136, 140) | <.0001 |
| Diastolic | 79 (77, 80) | 82 (80, 83) | 83 (82, 85) | 85 (84, 86) | <.0001 |
| Waist circumference, cm |  |  |  |  |  |
| Waist to hip ratio | 0.88 (0.87, 0.88) | 0.91 (0.91, 0.92) | 0.94 (0.94, 0.95) | 0.99 (0.98, 0.99) | <.0001 |
| Height, cm | 161.6 (160.5, 162.6) | 162.5 (161.5, 163.6) | 162.8 (161.7, 163.8) | 162.7 (161.6, 163.7) | <0.001 |
| Body fat percent | 19.1 (18.5, 19.8) | 23.5 (22.8, 24.1) | 26.5 (25.9, 27.1) | 30.8 (30.1, 31.4) | <.0001 |
| **Cardiac MR Variables** |  |  |  |  |  |
| LV ejection fraction, % | 62.1 (61.5, 62.6) | 62.7 (62.2, 63.2) | 63.2 (62.6, 63.7) | 63.2 (62.7, 63.8) | <0.001 |
| LV stroke volume index to body surface area, ml/m<sup>2</sup> | 47.6 (46.5, 48.7) | 45.2 (44.1, 46.3) | 43.0 (41.9, 44.1) | 40.6 (39.5, 41.8) | <.0001 |
| LV end diastolic volume index to body surface area, ml/m<sup>2</sup> | 76.9 (75.2, 78.5) | 72.3 (70.7, 74.0) | 68.2 (66.6, 69.9) | 64.6 (63.0, 66.3) | <.0001 |
| LV end systolic volume index to body surface area, ml/m<sup>2</sup> | 29.3 (28.6, 30.0) | 27.1 (26.5, 27.8) | 25.3 (24.6, 25.9) | 24.0 (23.4, 24.7) | <.0001 |
|  | Q1 | Q2 | Q3 | Q4 | P<sub>trend</sub> |
| LV myocardial mass systolic index to body surface area, g/m<sup>2</sup> | 63.1 (61.5, 64.8) | 62.1 (60.5, 63.7) | 61.7 (60.1, 63.3) | 61.1 (59.4, 62.7) | <0.001 |
| LV mass to volume ratio*, g/mL | 0.84 (0.81, 0.87) | 0.88 (0.84, 0.91) | 0.93 (0.89, 0.96) | 0.97 (0.94, 1.00) | <.0001 |

**Table 4** presents the association of VAT with LVMV (index of LV concentricity) in a multivariable model that includes sex, age, ethnicity, height, and CV risk factors (as measured by the IHRS). BSA was removed from the multivariable model due multicollinearity with VAT. For every 1 standard deviation increase in VAT, the LVMV increased by 0.037 g/mL (95% CI:0.032-0.041); and for every 5 unit increase in the IHRS, the LVMV increased by 0.024 g/mL (95% CI: 0.020-0.028). Notably, the association was consistent across both sexes, evidenced by the non-significance of the (VAT*sex) interaction (data not shown) and review of sex stratified analyses.

**Table 4:** Association of visceral adipose tissue with left ventricular mass-to-volume ratio (LVMV). The VAT variable represents a sex-specific standardized VAT, determined from [(VAT-mean)/sex-specific standard deviation]. Estimate from generalized linear mixed models with ethnicity (not shown) as a fixed effect and centre as a random intercept.

| OVERALL (N=6522) | MV Ratio - Univariable |  |  | MV Ratio - Multivariable |  |  |
| --- | --- | --- | --- | --- | --- | --- |
| Variable | Effect | 95% CI | P-value | Effect | 95% CI | P-value |
| VAT | 0.052 | 0.048 to 0.057 | <.0001 | 0.037 | 0.032 to 0.041 | <.0001 |
| Male (vs Female) | 0.122 | 0.114 to 0.131 | <.0001 | 0.124 | 0.112 to 0.135 | <.0001 |
| Age (by 10 years) | 0.042 | 0.037 to 0.047 | <.0001 | 0.020 | 0.016 to 0.025 | <.0001 |
| Height (per 10 cm) | 0.036 | 0.031 to 0.040 | <.0001 | -0.013 | -0.019 to -0.007 | <.0001 |
| INTERHEART Risk score (per 5-unit) | 0.053 | 0.049 to 0.057 | <.0001 | 0.024 | 0.020 to 0.028 | <.0001 |

### Role of Visceral Adiposity and Aging

Adjusted for height and IHRS, higher age was associated with higher LVMV. Specifically, each additional year of age was associated with a 0.0020 g/mL rise in LVMV (95% CI 0.0016 to 0.0025). Independent of other risk factors included in the model, the estimated impact of every 1 standard deviation increase in VAT on the LVMV is equivalent to 18.5 years of aging [Beta of VAT/(Beta of Age/10)]. This association was consistent across males and females, as indicated by the non-significant (age*sex) interaction (model not shown).

## Discussion

In this large population-based study of nearly 7000 generally healthy participants, we demonstrate that higher CV risk factor scores correlate with elevated VAT. Yet, even when accounting for CV risk factors, height, and age, an increase in VAT consistently corresponds to a rise in the LVMV across both males and females. Strikingly, a 1 standard deviation increase in VAT is associated with a LVMV that is 20 times higher than what is observed with natural aging alone and 1.5 times higher than with an integrated CV risk score. Future studies should specifically explore how interventions aimed at reducing visceral adiposity (VAT) influences cardiac remodelling.

### Role of Visceral Adipose Tissue in Cardiovascular Disease

VAT is increasingly recognized as a novel marker of inflammation associated with increased CVD.^19^ It is considered a highly active metabolic and endocrine organ that secretes cytokines and bioactive mediators (i.e., adiponectin) that can impact multiple pathways related to weight homeostasis, insulin sensitivity, inflammation, and lipid metabolism.^19^ The presence of VAT, which has distinct metabolic properties from subcutaneous adipose tissue, is strongly correlated with CV risk factors, such as elevated glucose levels, blood pressure, and atherogenic lipoprotein levels.^20,21^ This was supported by our data, as there was a higher burden of CV risk factors (i.e. diabetes, hypertension, waist-to-hip ratio) with each increasing quartile of MRI-detected VAT. Increased VAT accumulates from the relative inability of subcutaneous adipose tissue to expand through hyperplasia during surpluses in energy, resulting in lipid spillover and ectopic fat deposition in normally lean tissues of the heart, liver, skeletal muscle, pancreas, and kidney.^22^ Furthermore, excess VAT leads to increased production of circulating pro-inflammatory proteins associated with increased CVD.^22^

Using MRI, accurate quantification of VAT volume can be performed,^23^ which reflects the adipose tissue stored within the abdominal cavity. In our study, we demonstrate the independent association of VAT with LV concentricity, defined using CMR-derived LVMV, in both females and males. The alterations in LVMV is driven predominantly by a decrease in LVEDV, suggesting an increased burden of interstitial fibrosis leading to a “shrinking heart” or stiffer LV, which has been previously described using data from the MESA cohort.^23^ Strikingly, comparing normal references of LVEDV of males and females from our previously published data,^6^ we observe that for individuals with the highest quartiles of VAT, the mean LVEDV indexed to BSA for males and females (mean age 57 and 58 years, respectively), is comparable to that of participants nearly 10 years older in the 65 to 75 age group (69 ± 13 ml/m^2^ and 62 ± 9 ml/m^2^, respectively). This suggests VAT accumulation is related to accelerated cardiac aging that may be mediated by subclinical fibrosis.

### Role of Visceral Adiposity and Accelerated Cardiac Aging

Aging is a natural process that has been demonstrated to influence various physiological systems, including the cardiovascular system.^24^ One of the manifestations of aging on the heart is cardiac remodeling. Previous research has indicated that with every year increase in age, there is a small but measurable impact on LVMV.^3^ Specifically, LVMV increases by approximately 0.005g/mL for every year of age.^3^ This is in line with our own findings which indicate a change of 0.002g/mL per year of age.

However, when one factors in visceral adiposity, the scenario becomes more complex. Our data suggests that the presence of visceral adiposity can lead to an increase in LVMV that is equivalent to nearly 20 times the influence of aging alone. Additionally, the association of VAT with LVMV, which was independent of the CV risk score, further suggests that there are potentially specific, unknown pathways linking VAT to LVMV that are not mediated by vascular risk factors as captured by the IHRS. This finding lends support to the endocrine-inflammatory theory, which posits that VAT can release certain hormones or inflammatory factors that directly impact cardiac structures and functions.^25,26^

### Strengths and Limitations

This study has a number of strengths and some limitations. Strengths include the large size of the study and a precise measures of both VAT and cardiac contours on MRI images. The cross-sectional nature of the study limits our ability to make causal inferences. Furthermore, some of the cardio-metabolic risk factors were self-reported (e.g. type 2 diabetes), which may lead to an underestimation of CV risk factors, and therefore a possible overestimation of the effect size of VAT on LVMV.

Additionally, while the association between VAT and LVMV remains consistent across both sexes, the metabolic and inflammatory implications of VAT might be influenced by sex-specific factors, including hormonal profiles, adipocyte characteristics, and genetic predispositions that was not explored in this paper.^27,28^

## Conclusions

Among participants without clinical CVD in a large population of nearly 7000 individuals, we demonstrate a significant association of VAT and subclinical LV remodelling in males and females, represented by an increase in LVMV, independent of the effects of aging and other CV risk factors. We posit that deposition of VAT contributes to accelerated cardiac aging in both males and females.

## Data Availability

The datasets generated during and/or analyzed during the current study are available from the corresponding author on reasonable request.

## Declarations

Ethics approval and consent to participate

Research ethics approval was granted by the Hamilton Integrated Research Ethics Board, with consent obtained at each collaborating site as per site-specific regulations prior to participation in the study.

## Consent for publication

Not applicable.

## Competing interests

The University Hospital Zurich (CG) holds a research contract with GE Healthcare. Matthias G. Friedrich is board member, shareholder, and consultant of Circle Cardiovascular Imaging Inc.

## Funding

CAHHM was funded by the Canadian Partnership Against Cancer (CPAC), Heart and Stroke Foundation of Canada (HSF-Canada), and the Canadian Institutes of Health Research (CIHR). Financial contributions were also received from the Population Health Research Institute and CIHR Foundation Grant no. FDN-143255 to S.S.A.; FDN-143313 to J.V.T.; and FDN 154317 to E.E.S. In-kind contributions from A.R.M. and S.E.B. from Sunnybrook Hospital, Toronto for MRI reading costs, and Bayer AG for provision of IV contrast. The Canadian Partnership for Tomorrow Project is funded by the Canadian Partnership Against Cancer, BC Cancer Foundation, Genome Quebec, Ontario Institute for Cancer Research and Alberta Health and the Alberta Cancer Prevention Legacy Fund, Alberta Cancer Foundation. The PURE Study was funded by multiple sources. The Montreal Heart Institute Biobank is funded by Mr André Desmarais and Mrs France Chrétien-Desmarais and the Montreal Heart Institute Foundation. S.S.A. was supported by a Tier 1 Canada Research Chair in Ethnicity and Cardiovascular Disease (#CRC-2017-00024) and Heart and Stroke Foundation Chair in Population Health. P.A. was supported by a Ministry of Research and Innovation of Ontario Investigator Award. S.E.B. was supported by the Hurvitz Brain Sciences Research Program, Sunnybrook Research Institute, and the Department of Medicine, Sunnybrook Health Sciences Centre, University of Toronto. E.L. was supported by the Laval University Chair of Research & Innovation in Cardiovascular Imaging and the Fonds de recherche du Québec—Santé. J.-C.T. holds the Tier 1 Canada Research Chair in translational and personalized medicine and the Université de Montréal Pfizer endowed research chair in atherosclerosis. Some of the data used in this research were made available by the Canadian Partnership for Tomorrow Project along with BC Generations Project, Alberta’s tomorrow Project, Ontario Health Study, CARTaGENE, and the Atlantic PATH. Data were harmonized by Maelstrom and access policies and procedures were developed by the Centre of Genomics and Policy in collaboration with the Cohorts. Dr. Anand holds the Heart and Stroke Foundation Michael G DeGroote Chair in Population Health and a Canada Research Chair in Ethnic Diversity and Cardiovascular Disease. PURE-Mind Funding: The PURE study is an investigator-initiated study that was funded by the Population Health Research Institute, Hamilton Health Sciences Research Institute, the Canadian Institutes of Health Research, Heart and Stroke Foundation of Ontario, support from Canadian Institutes of Health Research’s Strategy for Patient Oriented Research, through the Ontario SPOR Support Unit, as well as the Ontario Ministry of Health and Long-Term Care and through unrestricted grants from several pharmaceutical companies (with major contributions from AstraZeneca [Canada], Sanofi-Aventis [France and Canada], Boehringer Ingelheim [Germany and Canada], Servier, and GlaxoSmithKline) and additional contributions from Novartis and King Pharma and from various national or local organizations in participating countries. In Canada, there was additional support for the PURE study from an unrestricted grant from Dairy Farmers of Canada and the National Dairy Council (US), Public Health Agency of Canada, and Champlain Cardiovascular Disease Prevention Network. The PURE Poland substudy was partially funded by grant 290/W-PURE/2008/0 from the Polish Ministry of Science and Higher Education, and the MRI substudy was supported by grant NCN 2015/17/B/NZ7-02963 from the Polish National Science Centre.

CG is supported by grants from the Swiss National Science Foundation (SNSF, # PP00P3_163892 and # PP00P3_190074), the Olga Mayenfisch Foundation, Switzerland, the OPO Foundation, Switzerland, the Novartis Foundation, Switzerland, the Swissheart Foundation, the Helmut Horten Foundation, Switzerland, the University Hospital Zurich (USZ) Foundation, the Iten-Kohaut Foundation, Switzerland, and the EMDO Foundation, Switzerland. Dr. Anand holds the Heart and Stroke Foundation Michael G DeGroote Chair in Population Health and a Canada Research Chair in Ethnic Diversity and Cardiovascular Disease.

## Role of the Funder/Sponsor

The funding sources had no role in the design and conduct of the study; collection, management, analysis, and interpretation of the data; preparation, review, or approval of the manuscript; and decision to submit the manuscript for publication.

## Authors’ contributions

J. Luu and C. Gebhard contributed to all aspects of the manuscript. D. Desai contributed to conception, design, data acquisition, analysis, interpretation, and revising of the manuscript. C. Ramasundarahettige and K. Schulze contributed to data analysis and interpretation, and manuscript revision. P. Awadalla and T. Dummer contributed to data acquisition and interpretation, and manuscript revision. F. Marcotte, J. Hicks, G. Lettre, V. Ho, and E. Larose contributed to the design, data acquisition and interpretation, and manuscript revision. A. Moody, E. Smith, J.C. Tardif, K. Teo, J. Vena, D. Lee, S. Anand, and M. Friedrich contributed to the design, data acquisition, and manuscript revision. All authors read and approved the final manuscript.

## Acknowledgement

In the preparation of this manuscript, we utilized OpenAI’s ChatGPT to assist with grammar corrections and rewording of material originally written by the authors, which constituted less than 1% of the entire document. We confirm that the substantive content, underlying research, and primary conclusions were solely the work of the authors and were not influenced or generated by ChatGPT.

## Additional Information

The Canadian Alliance of Healthy Hearts and Minds (CAHHM) investigators include the following individuals: Sonia S. Anand, MD, PhD, Department of Medicine, Department of Health Research Methods, Evidence, and Impact, McMaster University, Population Health Research Institute, Hamilton Health Sciences, Hamilton, Canada; Philip Awadalla, PhD, Department of Molecular Genetics, Ontario Institute for Cancer Research, University of Toronto, Toronto, Canada; Sandra E. Black, MA, MD, OC, Department of Medicine (Neurology), Sunnybrook Health Sciences Centre, University of Toronto, Toronto, Canada; Broët Philippe, MD, PhD, Department of Preventive and Social Medicine, École de santé publique, Université de Montréal, and Research Centre, CHU Sainte Justine, Montréal, Canada; Alexander Dick, MD, Department of Medicine, University of Ottawa Heart Institute, Ottawa, Canada; Trevor Dummer, PhD, MSc, Department of Epidemiology, Biostatistics, and Public HealthPractice, School of Population and Public Health, University of British Columbia, and BC Cancer Agency, Vancouver, Canada; Matthias G. Friedrich, MD, Department of Cardiology, McGill University, Montréal, Canada; Jason Hicks, MSc, Atlantic Partnership for Tomorrow’s Health, Dalhousie University, Halifax, Canada; David Kelton, MD, RPVI, Department of Medicine, William Osler Health System; Anish Kirpalani, MD, MASc, Department of Medical Imaging, St. Michael’s Hospital, University of Toronto, Toronto, Canada; Maria Bartha-Knoppers, PhD, Centre of Genomics and Policy, McGill University, Montréal, Canada; Scott A. Lear, PhD, Department of Pathology, Simon Fraser University, Burnaby, Canada; Eric Larose, DVM, MD, Department of Medicine, University of Laval, Quebec City, Canada; Russell J. de Souza, RD, ScD, Department of Health Research Methods, Evidence, and Impact, McMaster University, Hamilton, Canada; Douglas S. Lee, MD, PhD, ICES Central, Cardiovascular Research Program, Institute for Clinical Evaluative Sciences, Peter Munk Cardiac Centre University Health Network, Department of Medicine, University of Toronto, Toronto, Canada; Jonathan Leipsic, MD, Department of Radiology, University of British Columbia, Vancouver, Canada; Francois Marcotte, MD, Department of Cardiology, Montreal Heart Institute, University of Montreal, Montréal, Canada; Alan R. Moody, MBBS, Department of Medical Imaging, University of Toronto, Sunnybrook Health Sciences Centre, Toronto, Canada; Michael D. Noseworthy, PhD, PEng, Department of Electrical and Computer Engineering, McMaster University, St. Joseph’s Health Care, Hamilton, Canada; Grace Parraga, PhD, Department of Medical Biophysics, Robarts Research Institute, Western University, London, Canada; Louise Parker, PhD, Atlantic Partnership for Tomorrow’s Health, Dalhousie University, Halifax, Canada; Paul Poirier, MD, PhD, Institut de cardiologie et de pneumologie de Quebec, Université of Laval, Quebec City, Canada; Eric E. Smith, MD, MPH, Hotchkiss Brain Institute, Department of Clinical Neurosciences, University of Calgary, Calgary, Canada; Jean-Claude Tardif, MD, Department of Cardiology, Montreal Heart Institute, University of Montreal, Montréal, Canada; Koon K. Teo, MBBCh, PhD, Department of Medicine, Department of Health Research Methods, Evidence, and Impact, McMaster University, Population Health Research Institute, Hamilton Health Sciences, Hamilton, Canada; Jack V. Tu, MD, PhD, MSc, Department of Medicine, University of Toronto, Institute for Clinical Evaluative Sciences, Sunnybrook Schulich Heart Centre, Toronto, Canada (deceased); Jennifer Vena, PhD, Cancer Research and Analytics, Cancer Control Alberta, Alberta Health Services, Edmonton, Canada. The PURE-MIND investigators include the following individuals: Eric E. Smith, MD, MPH, Hotchkiss Brain Institute, Department of Clinical Neurosciences, University of Calgary, Calgary, Canada; Koon K. Teo, MBBCh, PhD, Department of Medicine, Department of Health Research Methods, Evidence, and Impact, McMaster University, Population Health Research Institute, Hamilton Health Sciences, Hamilton, Canada; Salim Yusuf, DPhil, Population Health Research Institute, Department of Medicine, McMaster University, Hamilton Health Sciences, Hamilton, Canada; Martin J. O’Donnell, MD, PhD, Population Health Research Institute, Department of Medicine, McMaster University, Hamilton Health Sciences, Hamilton, Canada; Michael D. Noseworthy, PhD, PEng, Department of Electrical and Computer Engineering, McMaster University, St. Joseph’s Health Care, Hamilton, Canada; Paul Poirier, MD, PhD, Institut de cardiologie et de pneumologie de Quebec, Université of Laval, Quebec City, Canada; Scott A. Lear, PhD, Faculty of Health Sciences, Simon Fraser University, St. Paul’s Hospital; Andreas Wielgosz, MD, PhD, University of Ottawa Heart Institute, Ottawa, Canada; Dorota Szcześniak, PhD, Department of Psychiatry, Division of Psychotherapy and Psychosomatic Medicine, Wroclaw Medical University, Wrocław, Poland; Andrzej Szuba, MD, PhD, Medicine, Department of Angiology, Hypertension and Diabetology, Wroclaw Medical University, Wrocław, Poland; Katarzyna Zatonska, MD, PhD, Department of Health Population, Wroclaw Medical University, Wrocław, Poland; Anna Zinny, MD, Department of General and Interventional Radiology and Neuroradiology, Wroclaw Medical University, Wrocław, Poland.

## Notes

### Competing Interest Statement

The authors have declared no competing interest.

### Author Declarations

This study complied with the Declaration of Helsinki, and each study site received institutional research ethics board approval for all procedures. All study participants provided written informed consent.

