## Supplementary figures and images for "Visceral Adiposity and Subclinical Left Ventricular Remodeling"

### Supplementary Figure 1

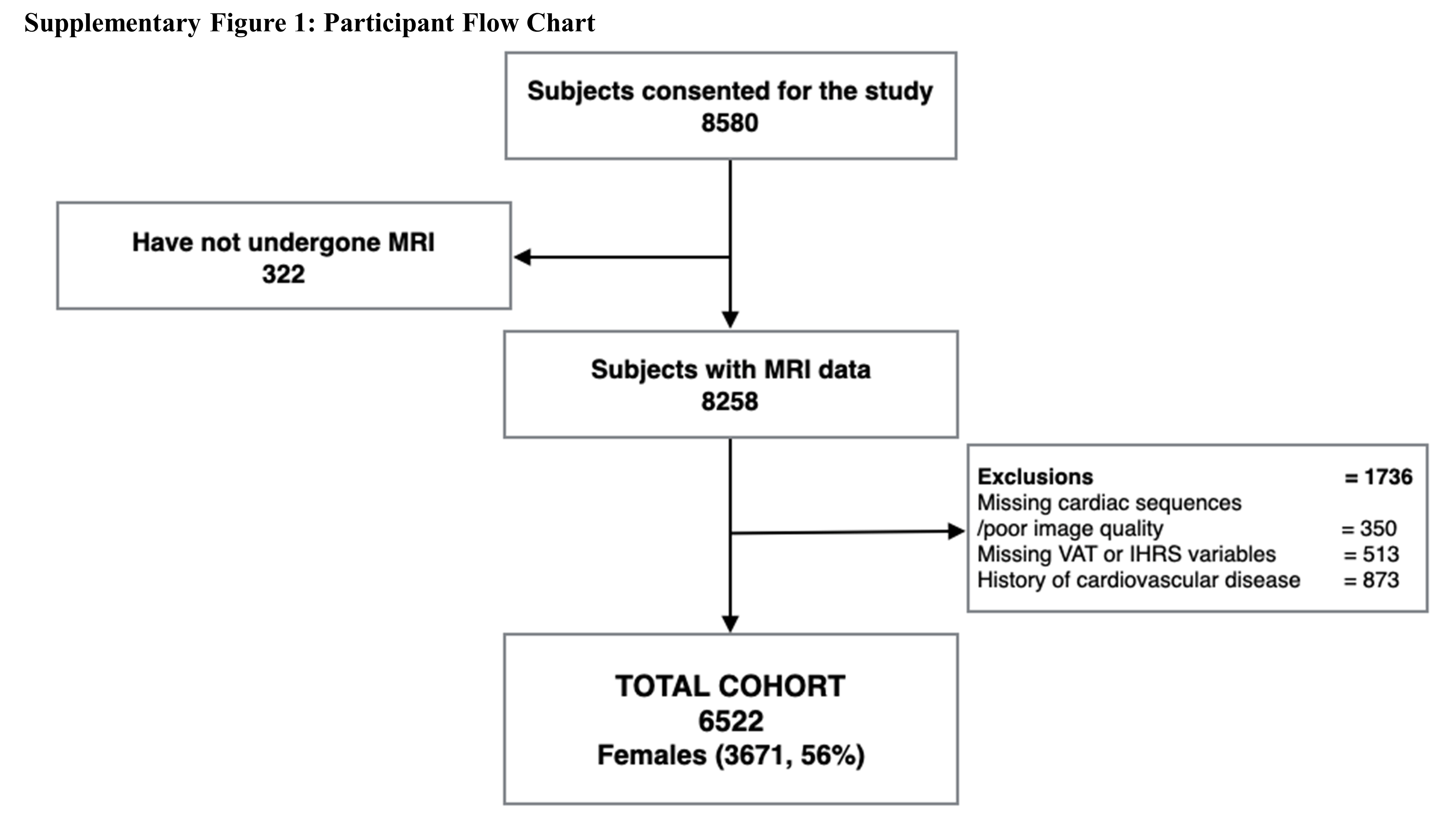
